# Longitudinal wastewater surveillance addressed public health priorities during the transition from “dynamic COVID-zero” to “opening up” in China: a population-based study

**DOI:** 10.1101/2023.03.25.23287563

**Authors:** Yinghui Li, Chen Du, Ziquan Lv, Fuxiang Wang, Liping Zhou, Yuejing Peng, Wending Li, Yulin Fu, Jiangteng Song, Chunyan Jia, Xin Zhang, Mujun Liu, Zimiao Wang, Bin Liu, Shulan Yan, Yuxiang Yang, Xueyun Li, Yong Zhang, Jianhui Yuan, Shikuan Xu, Miaoling Chen, Xiaolu Shi, Bo Peng, Qiongcheng Chen, Yaqun Qiu, Shuang Wu, Min Jiang, Miaomei Chen, Jinzhen Tang, Lei Wang, Lulu Hu, Chengsong Wan, Hongzhou Lu, Tong Zhang, Songzhe Fu, Xuan Zou, Qinghua Hu

**Author notes:** Corresponding authors: Hongzhou Lu, Tong Zhang, Songzhe Fu, Xuan Zou, Qinghua Hu. These authors contributed equally to the article (Yinghui Li, Chen Du, Ziquan Lv, Fuxiang Wang).

## Abstract

**Background:** Wastewater surveillance provides real-time, cost-effective monitoring of SARS-CoV-2 transmission. We developed the first city-level wastewater warning system in mainland China, located in Shenzhen. Our study aimed to reveal cryptic transmissions under the “dynamic COVID-zero” policy and characterize the dynamics of the infected population and variant prevalence, and then guide the allocation of medical resources during the transition to “opening up” in China.

**Methods:** In this population-based study, a total of 1,204 COVID-19 cases were enrolled to evaluate the contribution of Omicron variant-specific faecal shedding rates in wastewater. After that, wastewater samples from up to 334 sites distributed in communities and port areas in two districts of Shenzhen covering 1·74 million people were tested daily to evaluate the sensitivity and specificity of this approach, and were validated against daily SARS-CoV-2 screening. After the public health policy was switched to “opening up” in December 7, 2022, we conducted wastewater surveillance at wastewater treatment plants and pump stations covering 3·55 million people to estimate infected populations using model prediction and detect the relative abundance of SARS-CoV-2 lineages using wastewater sequencing.

**Findings:** In total, 82·4% of SARS-CoV-2 Omicron cases tested positive for faecal viral RNA within the first four days after the diagnosis, which was far more than the proportion of the ancestral variant. A total of 27,759 wastewater samples were detected from July 26 to November 30 in 2022, showing a sensitivity of 73·8% and a specificity of 99·8%. We further found that wastewater surveillance played roles in providing early warnings and revealing cryptic transmissions in two communities. Based on the above results, we employed a prediction model to monitor the daily number of infected individuals in Shenzhen during the transition to “opening up” in China, with over 80% of the population infected in both Futian District and Nanshan District. Notably, the prediction of the daily number of hospital admission was consistent with the actual number. Further sequencing revealed that the Omicron subvariant BA.5.2.48 accounted for the most abundant SARS-CoV-2 RNA in wastewater, and BF.7.14 and BA.5.2.49 ranked second and third, respectively, which was consistent with the clinical sequencing.

**Interpretation:** This study provides a scalable solution for wastewater surveillance of SARS-CoV-2 to provide real-time monitoring of the new variants, infected populations and facilitate the precise prediction of hospital admission. This novel framework could be a One Health system for the surveillance of other infectious and emerging pathogens with faecal shedding and antibiotic resistance genes in the future.

**Funding:** Sanming Project of Medicine in Shenzhen, Shenzhen Key Medical Discipline Construction Fund.

**Research in context:** *Evidence before this study:* We searched PubMed for articles published from December 1, 2019, to February 28, 2023, without any language restrictions, using the search terms “wastewater surveillance”, “SARS-CoV-2 shedding rate”, and “China”. After checking abstracts and full texts of the search results, we found that the field of wastewater-based epidemiology (WBE) has been considered as a powerful, rapid, and inexpensive tool to monitor SARS-CoV-2 transmission in recent years. Researchers realized that SARS-CoV-2 RNA in wastewater is mainly from the faecal virus shedding of infected individuals, and the number of infected individuals can be estimated using a prediction model based on the viral RNA load in wastewater and the faecal viral shedding rate. However, there are no published clinical data regarding the faecal shedding rates of the pandemic variant Omicron. In particular, no previous studies have reported the size of China’s SARS-CoV-2 infection after the public health policy was switched to “opening up” in December 7, 2022.

*Added value of this study:* This study highlights pioneering work in the use of wastewater surveillance of SARS-CoV-2 conducted during the transition from “dynamic COVID-zero” to “opening up” in China. The study reported first about the high proportion of faecal viral shedding of SARS-CoV-2 Omicron cases, showcasing the generality of wastewater surveillance for tracking Omicron prevalence. On the one hand, wastewater surveillance can play roles in providing early warnings and revealing cryptic transmissions and has the potential to replace city-wide nucleic acid screening under stringent control measures. On the flip side, wastewater surveillance allows for robust predictions of the number of infected individuals, the relative abundance of SARS-CoV-2 lineages, and the rate of hospital admission after the public health policy was switched to relaxed COVID-19 restrictions.

*Implications of all the available evidence:* Governments are in urgent need of a paradigm to shorten the time lag observed between recognition of a new emerging pathogen with the potential to cause the next pandemic (e.g., SARS-CoV-2) and the development of public health response (e.g., early warning, management and control of the communities, allocation of medical resources). Our findings suggest that the system developed in this study is not only a valuable epidemiological tool to accurately monitor the infection trend but also transforms wastewater surveillance into a public health management framework, which could be a One Health system for the surveillance of other infectious and emerging pathogens with faecal shedding and antibiotic resistance genes.

## Introduction

The limited population-based testing capacity of the SARS-CoV-2 pandemic triggered the wastewater-based epidemiology (WBE) method to assess COVID-19 prevalence. The theoretical basis of wastewater surveillance is that SARS-CoV-2 RNA in wastewater is mainly from the faecal virus shedding of infected persons. ^1-7^ To date, at least 72 countries and 288 universities and research institutes have carried out wastewater surveillance for SARS-CoV-2, which has played crucial roles in monitoring the infection trend and emerging variants, providing early warning signals for public health interventions.^8-13^

Recent studies have also shown the additional value of wastewater surveillance for the prediction of infected populations to guide public health actions. For instance, Ahmed et al. established a model to estimate the number of infected individuals based on the concentration of SARS-CoV-2 RNA, which was in agreement with clinical observations. ^14^ Fu et al. further considered the decay rate of SARS-CoV-2 RNA in the sewer system and proposed a revised model to reach an accurate perdition of infected persons. ^15^ The results from McMahan et al. also suggested that a modified susceptible-exposed-infectious-recovered (SEIR) model based on daily viral RNA load in wastewater provided rapid prevalence assessments of COVID-19 cases. ^16^

Since 2020, the “dynamic COVID-zero” strategy has been adopted in China for nearly three years. ^17^ The core of this strategy is to take timely measures to lockdown the communities for 7-14 days and quickly cut off the COVID-19 transmission chain. However, as the Omicron variant has become the dominant variant in the majority of countries in 2022 with a higher transmission capacity, the implementation of the “dynamic COVID-zero” strategy became unfeasible for most cities. Whether WBE would be a feasible solution to replace nucleic acid testing to monitor the infected population remains unknown. To this end, we first studied the faecal viral shedding rate of 1,204 COVID-19 cases in a designated hospital to evaluate the feasibility of wastewater surveillance for Omicron, and then we established a surveillance network in the Nanshan and Futian districts of Shenzhen City to evaluate the sensitivity and specificity of this approach. Afterward, since December 3, we conducted wastewater surveillance in wastewater treatment plants (WWTPs) and pump stations to monitor the dynamics of virus concentration and composition of variants. Finally, we adopted a predictive model to predict the number of infected persons during the transition from “dynamic COVID-zero” to “opening up”, ^14^ which provided timely information for policy makers and guided the allocation of medical resources to address this pandemic.

## Methods

### Study of faecal viral shedding of the Omicron variant

From August 9 to October 30, 2022, 1,204 infected individuals hospitalized in the Third People’s Hospital of Shenzhen, a designated hospital for COVID-19 cases in Shenzhen, were included in the study, and the hospital admission data were collected from electronic medical records of the hospital. Faecal samples were collected daily using sterilized cotton swabs and placed in SARS-CoV-2 inactivated sampling tubes. The GeneRotex96 automatic nucleic acid extraction platform (TIANLONG, China) was used to extract nucleic acids from samples, and the novel coronavirus 2019-nCoV nucleic acid detection kit (Hybribio, China) and ABI Prism7500 real-time PCR instrument (Hybribio, China) were used to perform RT-qPCR detection of SARS-CoV-2 RNA according to the manufacturer’s instructions. A positive faecal sample indicated that at least one target gene (*ORF1ab* or *N*) had a signal under the cut-off cycle threshold (Ct) value of 40. To unify the standard of quantification, the final viral concentration in faecal samples was calculated using the standard curve of *N*-carrying plasmids. Details about the quantitative analysis of viral loads in faeces are provided in the appendix.1 (p 1). Ethical approval for this study was provided by the ethical review board of Shenzhen CDC.

### Wastewater surveillance in port areas

Tentative wastewater surveillance was conducted in the districts of Futian and Nanshan of Shenzhen City. From July 26 to November 30, up to 334 sampling sites covering 1·74 million people were selected according to the potential risk of the infected persons imported from Hong Kong. ^18^ The average population covered by each sampling site was approximately 5,200 (50-70,000). Since December 3, we have conducted wastewater surveillance only at six WWTPs and nine pump stations in Futian District and Nanshan District. The population covered by each WWTP was approximately 592,000 (65,000-850,000), the population covered by each pump station was approximately 150,000 (10,000-572,000), and the total population was estimated to be approximately 3·55 million. Details about quantity changes, location, cover area, population and sewage flow of sampling sites are provided in the appendix.3-5 (pp 2-4).

From July 26 to November 30, wastewater was collected continuously from 8-11 am or 10-12 am based on the peak water consumption at each sampling point with 250 mL collected every 15 minutes, and 3 L of wastewater was collected in total. Since December 3, 2022, wastewater was collected continuously in 24 hours at each WWTP or pump station with 125 mL collected every hour, and 3 L of wastewater was collected in total. Wastewater samples were inactivated by a water bath at 60°C for 30 minutes and then enriched and concentrated by a modified “PEG precipitation method”.^19^ Nucleic acid extraction was performed using automatic nucleic acid extraction platform HBNP-9601A (Hybribio, China) or GENEDIAN EB1000 (DIAN Biotechnology, China). RT-qPCR detection and quantification were conducted as mentioned above. Details about the quantitative analysis of viral loads in wastewater are provided in the appendix.2 (p 1).

### Sequencing and analysis

Wastewater sequencing was performed on the BGI sequencing platform. Reverse transcription and amplification were performed using the ATOPlex RNA Multiplex PCR Amplification Kit V3.1 before library construction. DNA libraries were prepared using the MGIEasy Fast PCR-FREE digestion Library Preparation kit. Sequencing was completed on the DNBSEQ-G99 sequencing platform using the single-end sequencing 100 bp+ double-end barcode (SE100+10+10) strategy. The MGI MetargetCOVID software was used to complete the parametric assembly of SARS-CoV-2 sequences, and then the mutation branches and mutation sites of SARS-CoV-2 sequences were obtained using nextclade and Pangolin analysis websites. Single-nucleotide polymorphisms (SNPs) were identified using Snippy (https://github.com/tseemann/snippy) by mapping sequencing reads to the reference genome (MN908947.3, Wuhan-Hu-1). SNP distances between viral genomes were calculated based on concatenated SNPs using snp-dists (https://github.com/tseemann/snp-dists).^20^

To determine SARS-CoV-2 lineage abundance in wastewater, we first performed quality control, alignment and primer sequence trimming using MGI MetargetCOVID software. In brief, sequencing reads were aligned to the reference sequence (MN908947.3) with BWA mem, and reads with hard or soft clips >10 were filtered using the SAMclip method. The resulting bam files were used to calculate the sequencing depth and coverage by samtools, where samples with less than 60% genome coverage were excluded. We used the Freyja tool (v1.3.12) to recover the relative abundances of SARS-CoV-2 lineages from wastewater samples using lineage-defining mutational “barcodes” derived from the UShER global phylogenetic tree (as of January 29, 2023). ^21^ Lineages were excluded if they either had overall low abundance (<1%) or showed low minor allele frequency (<1%) or sequencing depth (<10X) in their defining mutations.

Clinical sequencing data of 478 human oropharyngeal swab samples were collected from “dynamic COVID-zero” case report before December 6 and COVID-19 integrated surveillance system after December 7, which were published by Health Commission of Shenzhen Municipality.

### Prediction of infected persons based on WBE

We first calculated the decay rate of SARS-CoV-2 RNA in a wastewater pipeline by the following equation:

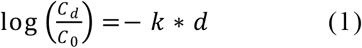

where C_0_ and C_d_ are the SARS-CoV-2 RNA concentrations in the wastewater at travel distances of 0 and d km, respectively. *d* is the distance from a community to a WWTP, and *k* is the decay rate constant, which is 0.12 in winter, as suggested by our previous study. ^15^ As C_d_ was obtained by qPCR assay in the WWTP, we then used Eq. (1) to obtain C_0,_ which is the corrected SARS-CoV-2 RNA concentration.

Afterwards, the daily number of actively infected persons in the catchment of a WWTP can be estimated on the basis of the number of virus copies present in sewage (numerator) and the amount of virus shedding per person per day (denominator) by the following equations (2):

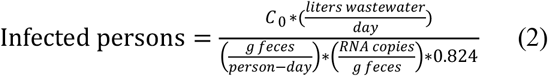

where 0.824 is the shedding rate of the infected population as determined by the previous section.

In the above equation, the amount of SARS-CoV-2 RNA copies/g in feces per person was modelled as a log-uniform distribution ranging from 4.2 to 9.67 log_10_ copies/g as determined by the previous section. The amount of faecal production per person was set at 128 g per day. Based on Equation (2), the Monte Carlo simulation was conducted with ModelRisk version 6.0. The number of iterations per simulation was 10,000. The predicted infection cases are reported as the median and 95% confidence interval (CI) determined by bootstrapping the model with 100 experiments.

### Hospitalization data collection in Nanshan district

We obtained medical records and compiled data for hospitalized patients with laboratory-confirmed COVID-19 in Nanshan district, as reported to the Health Commission of Shenzhen Municipality between December 12, 2022, and January 10, 2023.

### Data analysis

The collection and testing of faecal and wastewater samples were recorded using Microsoft Excel version 2210. Significance analysis of faecal viral shedding proportion and load between participants were analysed using IBM SPSS Statistics 27. Wastewater data were plotted and statistically analysed using GraphPad Prism version 8.4.0 (GraphPad Software, La Jolla, CA, USA).

### Role of the funding source

The funders of the study had no role in study design, data collection, data analysis, data interpretation, or writing of the report.

## Results

### Faecal viral shedding dynamics of SARS-CoV-2 Omicron cases

A total of 1,204 COVID-19 cases were enrolled to analyse the dynamics of faecal viral shedding. Viral variants were identified in 80·2% of participants and were all classified as Omicron, mainly including BA.5.2, BF.7 and BA.5.2.1 (Table 1). As shown in the appendix.6 (p 5), 63·6% (171/269) of participants tested positive for faecal SARS-CoV-2 RNA on day 1 (the next day after the diagnosis), and the faecal viral shedding proportion gradually increased to 71·5% (447/625) on day 4 and then gradually declined to 52·4% (98/187) on day 7, 21·8% (212/973) in the second week and 10·3% (23/223) in the third week. As the faecal viral shedding proportion peaked on day 4, the period from day 0 to day 4 was defined as the acute period of faecal viral shedding, and 82·4% (833/1,011) of participants tested positive for faecal SARS-CoV-2 RNA during the acute period, indicating the utility of wastewater surveillance in reflecting the majority of the infected population. A decline in faecal viral RNA concentration was observed since enrolment in the study, particularly obvious from day 4 to day 7 (appendix.6 p 5). The faecal viral loads of participants during the acute period ranged from 1·6×10^4^ to 4·1×10^10^ copies/g with a mean value of 6·7×10^7^ copies/g.

**Table 1:** Characteristics of participants included between August 9 and October 30, 2022

| Participants (n=1204) |  |
| --- | --- |
| Sex |  |
| Male | 636 (52.8%) |
| Female | 568 (47.2%) |
| Age, years | 37.3 (16.2) |
| 0-9 | 78 (6.5%) |
| 10-19 | 83 (6.9%) |
| 20-29 | 209 (17.4%) |
| 30-39 | 297 (24.7%) |
| 40-49 | 247 (20.5%) |
| 50-59 | 203 (16.9%) |
| 60-69 | 66 (5.5%) |
| 70-79 | 15 (1.2%) |
| 80-91 | 6 (0.5%) |
| Subvariant of the virus |  |
| Omicron BA.5.2 | 710 (59.0%) |
| Omicron BF.7 | 70 (5.8%) |
| Omicron BA.5.2.1 | 69 (5.7%) |
| Omicron BA.2.2 | 37 (3.1%) |
| Omicron BA.2.76 | 29 (2.4%) |
| Omicron BF.15 | 18 (1.5%) |
| Omicron BA.5.1 | 10 (0.8%) |
| Omicron BA.5.6 | 8 (0.7%) |
| Omicron BA.2.12.1 | 5 (0.4%) |
| Omicron BA.4.1 | 5 (0.4%) |
| Omicron BA.2.3.11 | 1 (0.1%) |
| Omicron BA.2.36 | 1 (0.1%) |
| Omicron BA.5 | 1 (0.1%) |
| Omicron BA.5.3.1 | 1 (0.1%) |
| Omicron BE.3 | 1 (0.1%) |
| Non-sequencing | 238 (19.8%) |
| Clinical Symptom Category |  |
| Patients with mild symptoms | 733 (60.9%) |
| Asymptomatic cases | 471 (39.1%) |
| COVID-19 Vaccine Dose |  |
| 0 | 96 (8.0%) |
| 1 | 22 (1.8%) |
| 2 | 278 (23.1%) |
| 3 | 774 (64.3%) |
| 4 | 6 (0.5%) |
| Information loss | 28 (2.3%) |
| COVID-19 Vaccine Type |  |
| Inactivated vaccine | 1020 (84.7%) |
| Recombinant protein vaccine | 12 (1.0%) |
| Adenovirus vector vaccine | 11 (0.9%) |
| mRNA vaccine | 18 (1.5%) |
| Mix And match | 20 (1.7%) |
| Not vaccinated | 96 (8.0%) |
| Information loss | 27 (2.2%) |
| Hospital Day(HOD) | 14.0 (3.6) |
| 3-7 | 25 (2.1%) |
| 8-14 | 665 (55.2%) |
| 15-21 | 452 (37.5%) |
| 22-28 | 25 (2.1%) |
| >28 | 6 (0.5%) |
| Information loss | 31 (2.6%) |
| Source Of Case |  |
| Domestically transmitted case | 1061 (88.1%) |
| Imported case | 131 (10.9%) |
| Information loss | 12 (1.0%) |

Data are mean (SD) or n (%).

We also analysed factors influencing faecal viral shedding during the acute period, including subvariant of Omicron, sex, age, symptom and vaccination (appendix.7 p 6). The faecal viral shedding proportion of Omicron subvariant BA.2.76 (100%, 29/29) was higher than that of BA.5.2 (85·8%, 503/586), BA.5.2.1 (83·6%, 51/61) and BF.7 (75·4%, 46/61), while there was no significant difference between their faecal viral loads (p=0.2744). The faecal viral shedding proportion gradually increased with age. Notably, the proportion and viral load of faecal viral shedding of people aged 60 years and older was significantly greater than that of children under 9 years old (p=0.000914, p=0.0094, respectively). No significant difference (p>0.05) in the proportion and viral load of faecal viral shedding was found between females and males, between mild cases and asymptomatic carriers, or between participants inoculated with inactivated vaccine and participants unvaccinated, suggesting the generality of wastewater surveillance for different population groups.

### Tentative wastewater surveillance during the “dynamic COVID-zero” period

As faecal shedding occurred in the majority of the population, we conducted tentative wastewater surveillance in the Nanshan and Futian districts of Shenzhen City with up to 334 wastewater sampling sites to evaluate sensitivity and specificity and evaluate whether wastewater surveillance can help nucleic acid tests find missed infected persons (figure 1A). Measures including early warning, building-level tracing and wastewater sequencing were adopted after SARS-CoV-2 RNA was detected in wastewater (appendix.8 p 7).

**Figure 1:**
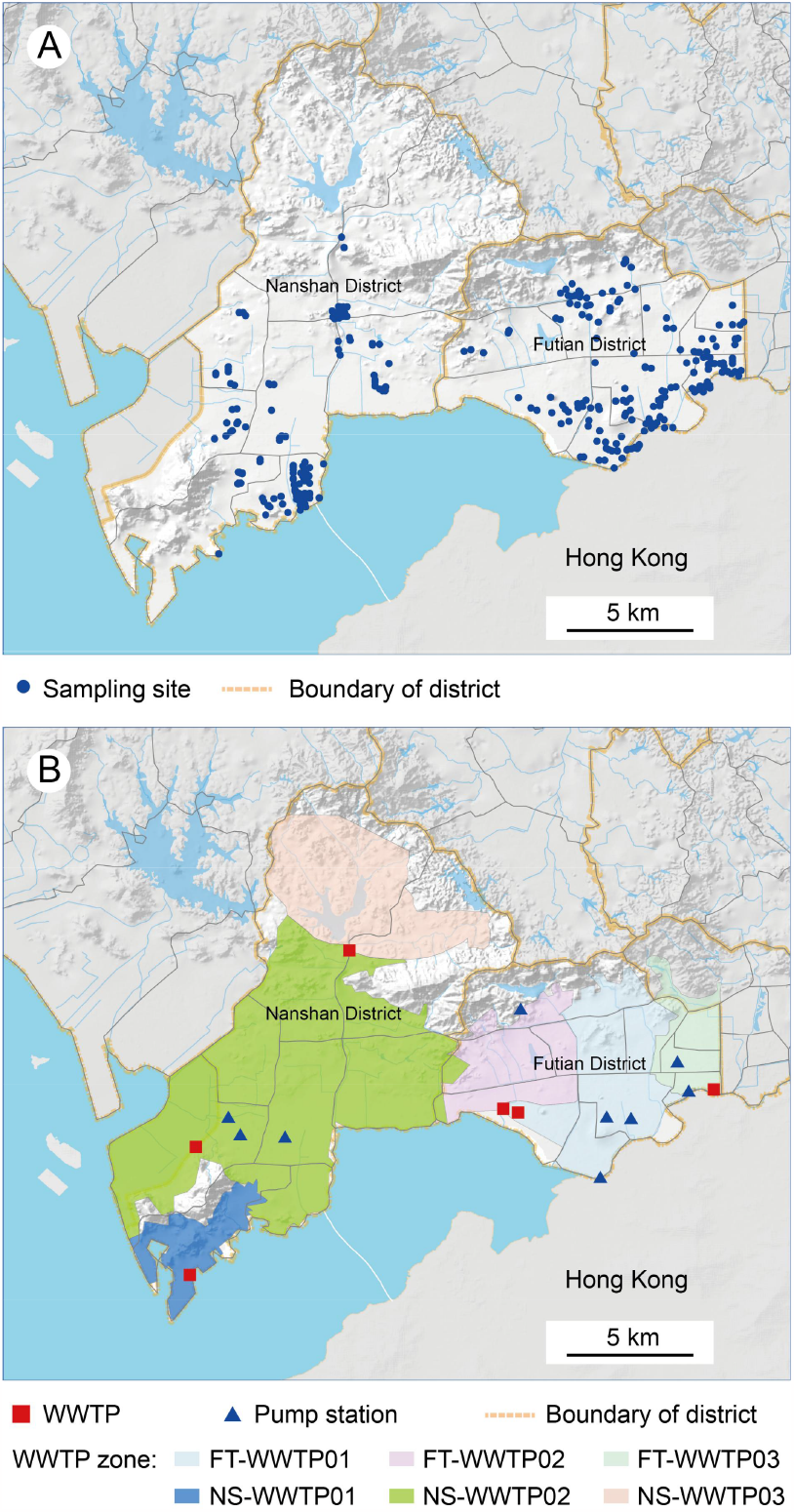
Location of sampling sites for SARS-CoV-2 detection. (A) During the “dynamic COVID-zero” period, from July 26 to November 30, 2022, up to 334 sampling sites were distributed in Futian District and Nanshan District. (B) After the switch of policy transmitted to “open-up”, from December 3, 2022 to January 20, 2023, wastewater surveillance was only conducted at 6 WWTPs and 9 pump stations in Futian District and Nanshan District.

From July 26 to November 30, 2022, 27,759 wastewater samples were collected, of which 239 tested positive. Very good agreement was observed between the number of positive sites detected by both wastewater surveillance and daily city-wide nucleic acid screening (figure 2A). Associated cases could be found in the coverage area of 77.8% (186/239) of sampling sites that tested positive, indicating that wastewater surveillance allows early warning of new cases in the coverage area. However, 26·2% (66/252) of the sampling sites that tested positive by daily city-wide nucleic acid screening were missed by wastewater surveillance, indicating that there was a certain proportion of missing inspection in wastewater surveillance, which might be caused by the fact that a fraction of infected individuals did not shed virus or shed virus beyond the time of wastewater sampling. Wastewater surveillance had a sensitivity of 73·8% and a specificity of 99·8% (figure 2B).

**Figure 2:**
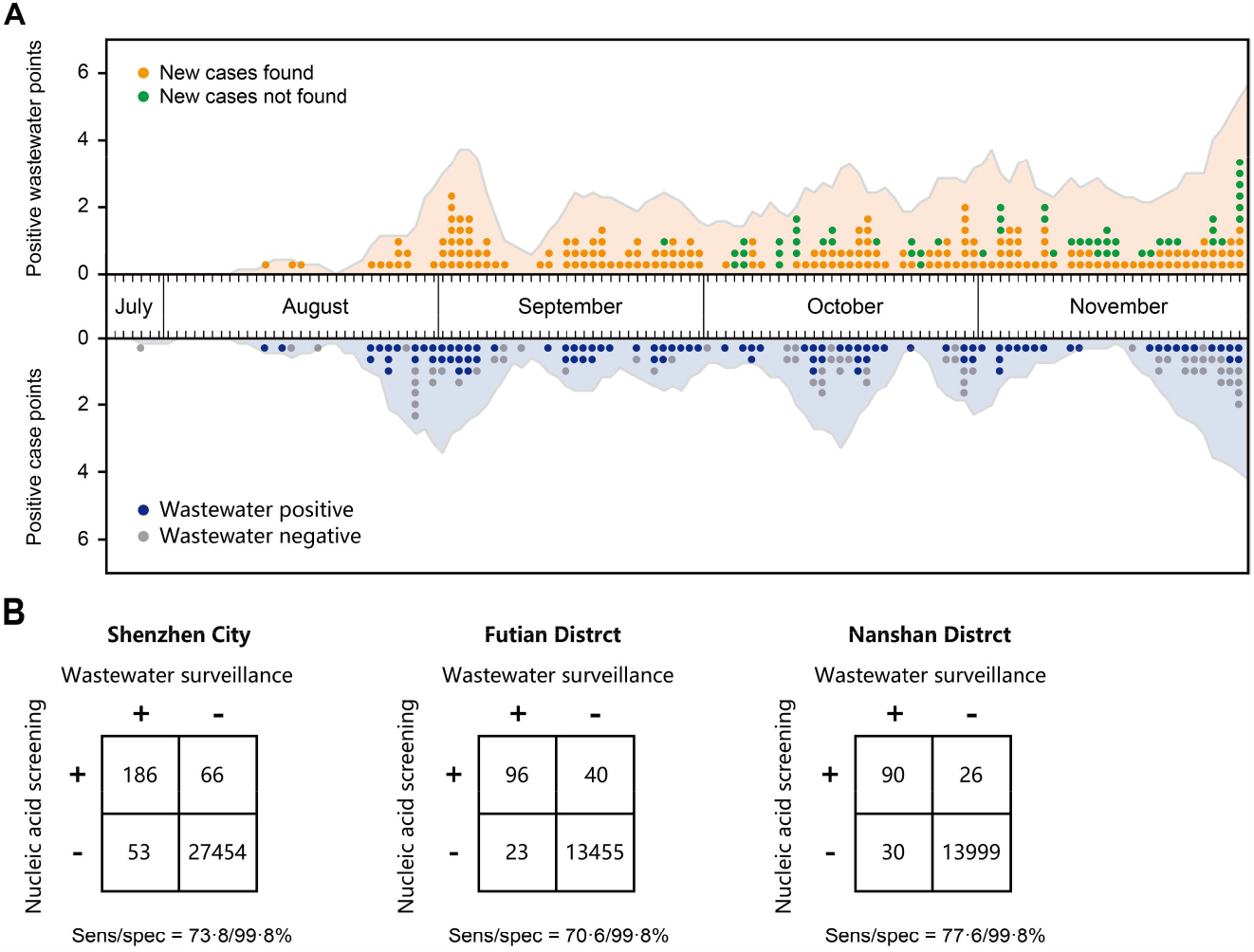
Wastewater surveillance compared with daily city-wide SARS-CoV-2 nucleic acid screening. (A) Reported number of positive sites found by wastewater surveillance and daily nucleic acid screening in Futian District and Nanshan District, and a 7-day trailing average (outline of the light yellow and light blue zone). (B) Contingency tables for wastewater surveillance and daily city-wide SARS-CoV-2 nucleic acid screening validation sets in Shenzhen City, Futian District, and Nanshan District, respectively.

Next, we found two cryptic transmission events that were missed by daily city-wide nucleic acid screening (appendix.9 p 8). From August 9 to September 21, 2022, routine wastewater surveillance did not detect SARS-CoV-2 circulation in the Badeng community until SARS-CoV-2 RNA was identified continuously in wastewater from September 22 to 24. Then, nucleic acid screening found two infected individuals on September 24th, with negative results on September 22nd and 23rd. Analysis of viral genomes revealed only one SNP difference between two infected individuals and the 23rd wastewater sample with a coverage of 93%, indicating that SARS-CoV-2 RNA in wastewater was from these two infected individuals. In another case, two outbreaks, NS1013 caused by Omicron subvariant BF.7 and NS1029 caused by BA.5.2.1, occurred in Nanshan community in October 2022, both of which were provided early warning through routine wastewater surveillance. However, SARS-CoV-2 RNA continued to be detected in wastewater in November and December, but no new infected individuals were found in the coverage area. These results demonstrate that wastewater surveillance has played roles in early warnings and revealing cryptic transmissions, suggesting that wastewater surveillance is an alternative to nucleic acid testing under stringent control measures.

### Wastewater surveillance during the transition to “opening up”

On December 7, 2022, authorities switched the public health policy from “dynamic COVID-Zero” to “opening up”. After that, we conducted wastewater surveillance only at the WWTP and pump station to assess the impact of policy change on the epidemic situation of COVID-19 in Shenzhen (figure 1B). Measured concentrations of SARS-CoV-2 RNA showed two waves of the COVID-19 pandemic on approximately December 20 and 27, respectively, and peaked on December 27 in both Futian and Nanshan districts (figure 3), suggesting that the COVID-19 pandemic in Shenzhen reached its peak on December 27.

**Figure 3:**
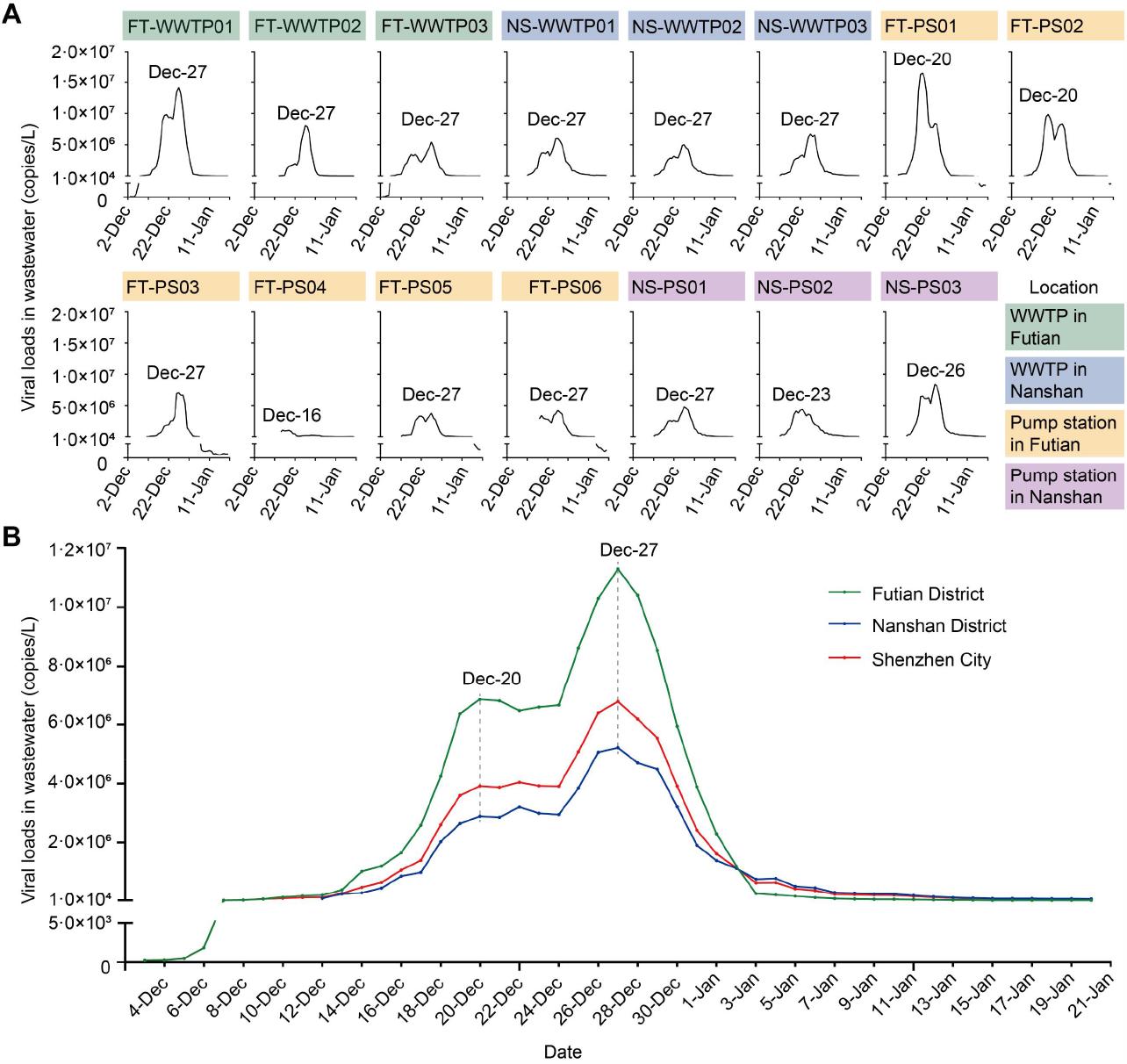
Wastewater-derived viral loads of SARS-CoV-2 in Shenzhen after the public health policy was switched to “open-up”. Viral loads of SARS-CoV-2 in wastewater (a 5-day trailing average) in 6 WWTPs and 9 pump stations in Futian District and Nanshan District. (B) Viral loads in wastewater (a weighted average of WWTPs) in Futian District, Nanshan District, and Shenzhen City (a weighted average of two districts).

Based on the concentration of SARS-CoV-2 RNA in wastewater and the average faecal viral shedding rate of Omicron for individuals, we first adopted Ahmed’s model to predict the daily number of existing infected individuals and cumulative infection rates in the Futian and Nanshan districts of Shenzhen from December 3, 2022, to January 20, 2023. Initially, we compared the prediction results based on wastewater data from a WWTP in Futian District (WWTP-FT03) with the infected persons detected by city-wide nucleic acid screening from December 3^rd^ to 7^th^ of 2022 (appendix.10 p 9). The results showed that the number of predicted persons was in line with the ones obtained from nucleic acid screening except for the data from December 7^th^ (city-wide nucleic acid screening was only partially implemented on that day). In Futian District, the first wave of the COVID-19 pandemic occurred on approximately December 20, 2022, when 291,395 cases were predicted. The second wave appeared around December 27, 2022, with 977,524 cases estimated (figure 4A). Likewise, the first wave in Nanshan District was also observed around December 20, which recorded 362,264 cases. The second wave was witnessed on December 28, with 411,785 predicted cases (figure 4B). On January 20, 2023, the cumulative rate of infections in Futian district was as high as 93·8%, while the cumulative rate of infections in Nanshan district was approximately 81·6%.

**Figure 4:**
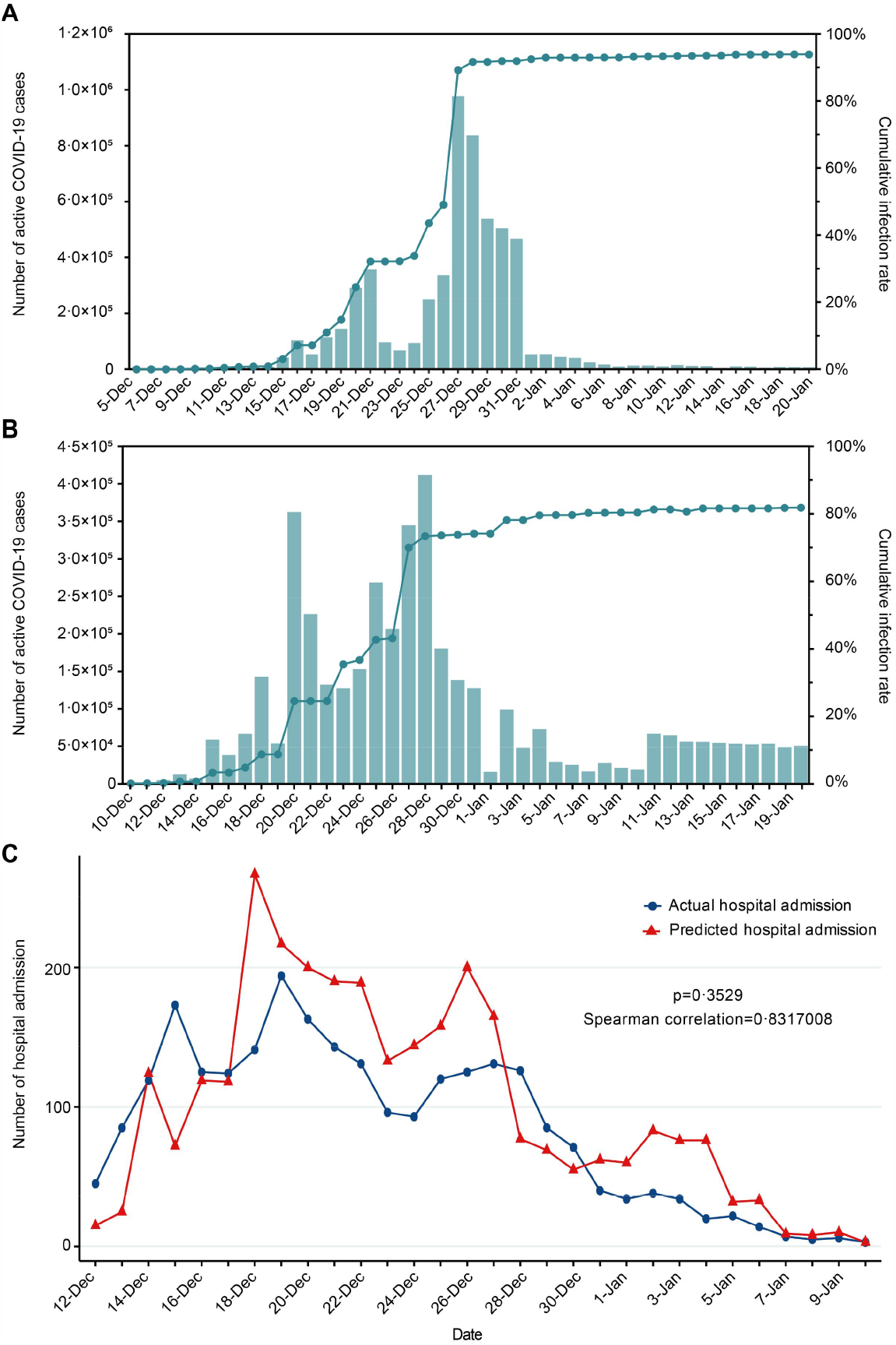
Daily predicted active COVID-19 cases in two districts and predicted number of daily hospital admission in Nanshan District. Daily predicted COVID-19 cases and cumulative infection rate in Futian (A) and Nanshan District based on wastewater data in Shenzhen, China (Dec 2022 - Jan 2023). Predicted values represented the median values of Monte Carol Simulation. 95% CI is not shown for clarity. (C) The predicted daily number of hospital admission vs actual daily number of hospital admission.

Furthermore, we adopted the research results from Jassat et al. and Nyberg et al. and estimated that 2·2% of elderly infected persons (age >65), 1% of children aged below 5 and 0·05% of remaining infected persons required hospital admission. ^22,23^ Thus, we calculated the daily number of hospital admissions based on wastewater data. Relative to the actual number of hospital admissions in Nanshan district, the prediction of hospital admission was slightly higher than the actual number with no significant difference, with a total of 2,815 predicted inpatients (figure 4C). In addition, the predicted peak of hospital admission was 1-2 days earlier than the actual peak, indicating the value of prediction for early warning and preparedness of medical resources.

To monitor the relative abundance of variants in wastewater, we selected 37 representative wastewater samples collected from December 13, 2022, to January 17, 2023, for sequencing. For comparison, we also analysed the clinical sequencing data of 478 human oropharyngeal swab samples collected from December 1, 2022, to January 13, 2023 (appendix.11 p 10). Wastewater sequencing revealed that 36 subvariants existed in wastewater, BA.5.2.48 accounted for the vast majority (62·4%), and BF.7.14 (24·6%) and BA.5.2.49 (5·6%) ranked second and third, respectively, which was consistent with clinical sequencing (figure 5). These subvariants were branches of BA.5.2 and BF.7, which were first detected in travelers from Hong Kong in July and October 2022, indicating the tremendous input risk of the port areas in Shenzhen adjacent to Hong Kong. Additionally, we found extra 29 variants with low abundance compared to clinical sequencing, indicating a low level of cryptic transmissions in Shenzhen. These results suggested a low degree of virus variation in the COVID-19 pandemic in Shenzhen during the transition from “dynamic COVID-zero” to “opening up” in China. No new emerging SARS-CoV-2 variants with the potential to cause the next pandemic (e.g., BQ.1.1, XBB.1.5 and CH.1.1) have been found by either wastewater sequencing or clinical sequencing efforts.

**Figure 5:**
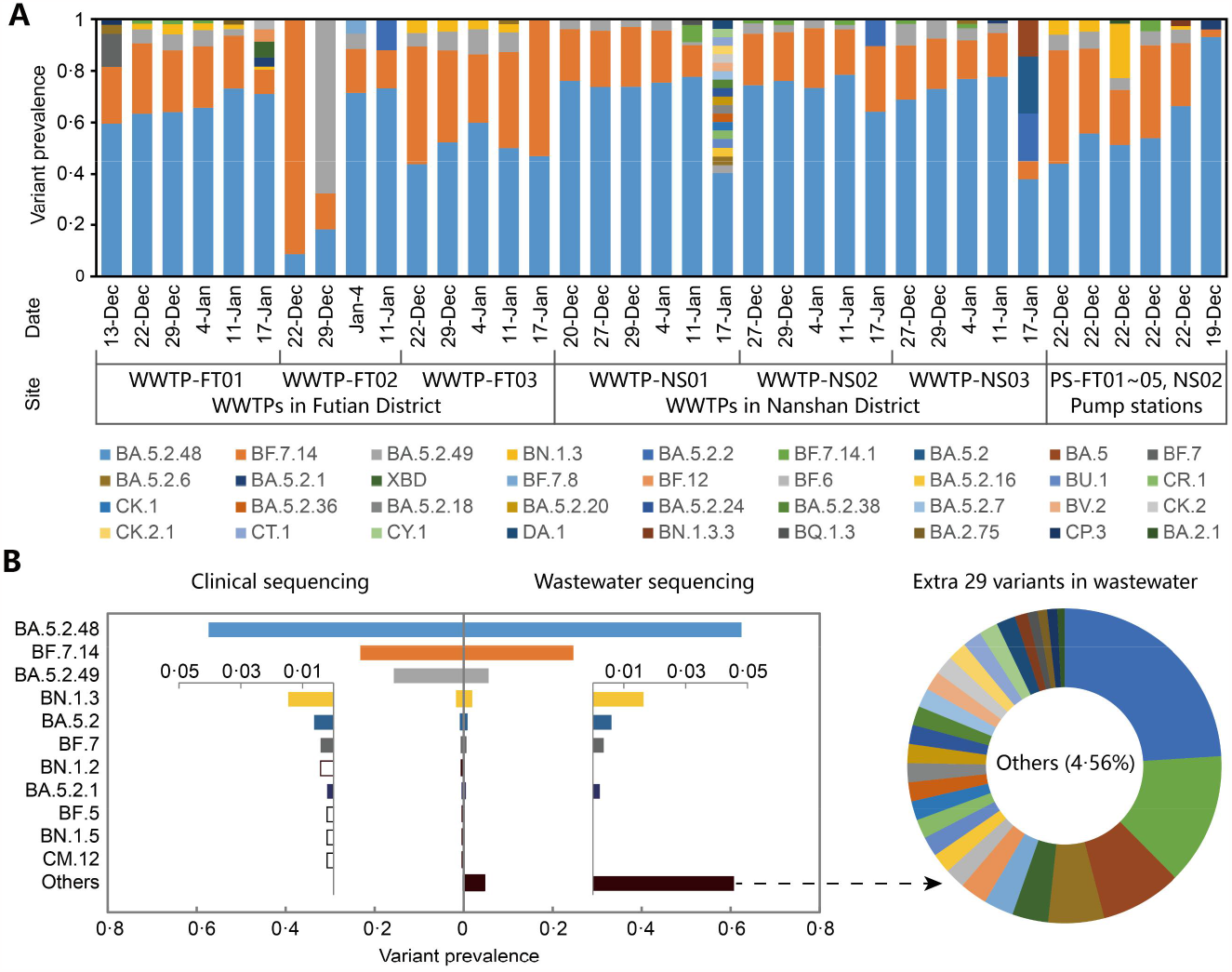
Variant prevalence of representative wastewater samples and human oropharyngeal swab samples. (A) Variant prevalence of representative wastewater samples (n=37). (B) Comparison of variant prevalence between wastewater samples and human oropharyngeal swab samples (n=478). Solid bars indicated variants that could be identified in wastewater and hollow bars indicated that variants were not be identified in wastewater but identified by clinical sequencing.

## Discussion

In this study, we first demonstrated that faecal viral shedding occurred in the majority of SARS-CoV-2 Omicron cases regardless of sex, symptoms and inoculation of inactivated vaccine, suggesting the generality of wastewater surveillance for tracking Omicron prevalence. After that, we employed WBE to monitor SARS-CoV-2 in up to 334 sampling sites and found that wastewater surveillance had a sensitivity of 73·8% and a specificity of 99·8%, contributing to the potential to replace nucleic acid tests. Based on the above findings, we adopted daily wastewater surveillance in 15 WWTP or pump station sites in two districts, which showed the robustness of wastewater surveillance in reflecting the infected population and the relative abundance of SARS-CoV-2 lineages after the switch of policy transmitted to “opening up”.

The previous studies on faecal viral shedding of COVID-19 cases mainly focused on the ancestral variant, and the proportion of faecal viral shedding of the ancestral variant was 21% to 59%.^24-30^ We conducted the study with a sample size of over 1,000 infected cases, which provided valuable information for the utility of wastewater surveillance. In total, 82·4% (n=1,011) of participants had samples collected during the acute period, indicating that wastewater surveillance by Omicron was a reliable approach to monitor infection trends. The range of faecal viral load of participants during the acute period was 1.6×10^4^ to 4.1×10^10^ copies/g with a mean value of 6.7×10^7^ copies/g, which was consistent with previous studies^1,31-33^, indicating little difference between SARS-CoV-2 variants.

City-wide SARS-CoV-2 nucleic acid screening has been an efficient tool in China to estimate the prevalence of infection, which could inform pandemic management. ^34^ However, the cost and its considerable inconvenience on people’s daily life and social functioning are unaffordable. In our study, relative to daily city-wide nucleic acid screening, wastewater surveillance provided a robust and affordable method for estimating the prevalence of infected persons in Shenzhen. It had a sensitivity of 73·8% and a specificity of 99·8% compared to daily city-wide nucleic acid screening, demonstrating the replaceability of this approach in reflecting the overall infection trends. Moreover, wastewater surveillance has played roles in early warning and revealing cryptic transmission in communities in Shenzhen. Therefore, wastewater surveillance could be used as an alternative method of population-based nucleic acid screening to produce actionable data for public health interventions for new emerging pathogens, such as SARS-CoV-2.

In this study, for any given sewershed, the numbers of infected cases were calculated and modelled by using the daily sewage flow rate multiplied by the virus RNA concentration divided by the RNA copies per person per day. The results showed that over 80% of the population was infected in both Futian District and Nanshan District. This is consistent with the public health data of Guangdong Province. ^35^ This approach has been widely used in previous studies with great success. ^14-16,36^ With the data of predicted infected cases and hospital admissions, policy makers now have a new valuable tool to tailor policy and make some critical decisions. For instance, the increase in predicted infected cases can serve as a leading indicator implying that active transmission is underway, even before new cases do not visit the hospitals. In particular, if a rapid increase in predicted cases is observed, public health authorities can predict hospital admission in the next few days and can be better prepared to arrange hospital beds and allocate medical resources. Additionally, once RNA levels decline sufficiently, wastewater surveillance can once again be used as a critical indicator of a resurgence of transmission.

Our study has three limitations. First, there was a lack of continuous faecal samples collected during the study of faecal viral shedding. Only faecal samples of 92 participants were collected daily from day 1 to day 4, and 93·5% of them tested positive for faecal SARS-CoV-2 RNA during the acute period. In addition, we conducted wastewater surveillance on December 12 in Nanshan District, which was five days later than December 7 (opening day) and might underestimate the infection rate in the first wave of the COVID-19 pandemic. Finally, the model predictions rely on several key parameters with considerable variations, including the distribution of shedding rates, temperature of the sewage, and decay rate of the RNA in the sewer system, resulting in high variability of the modelling results. Furthermore, this study did not encompass the effect of variants, which might slightly alter the model inputs. Further investigations are needed to improve this proposal. However, given the reliability of the modelling approach validated by previous studies, ^14-16^ we can safely reach the conclusion that SARS-CoV-2 quickly infected the majority of populations in the studied regions.

This study provided the first study of faecal viral shedding dynamics of Omicron, and tentative wastewater surveillance also demonstrated that wastewater surveillance has high sensitivity and specificity to replace nucleic acid tests for monitoring SARS-CoV-2. Based on these findings, we conducted wastewater surveillance in WWTPs and pump stations, monitored the infection trends and estimated the dynamics of populations in two border districts in Shenzhen. This is the first epidemiological description during the transition from “dynamic COVID-zero” to “opening up” in China. Predicted COVID-19 cases based on wastewater surveillance demonstrated that the majority of persons in the studied regions were infected, indicating a huge impact of policy change on the transmission of COVID-19. Our findings suggest that SARS-CoV-2 in wastewater also transforms wastewater epidemiology into a public health management framework, which could be a One Health system for the surveillance of other emerging pathogens with faecal viral shedding and antibiotic resistance genes in the future.

## Data Availability

All data needed to evaluate the conclusions in the Article are present in the main Article text and in the appendix. All raw sequencing data are available via the NCBI Sequence Read Archive under the BioProject ID PRJNA937922.

https://dataview.ncbi.nlm.nih.gov/object/PRJNA937922?reviewer=57rdc9outr3lv1mlq6q12b2kds

## Contributors

QH, YL, XZ, SF, TZ, HL, LZ designed, initiated and coordinated the study. YL, CD and ZL wrote the first draft. FW organized the collection of faecal samples. CD, ZL, SF, YP, WL and YF accessed and verified the data and made the tables and figures. JS, CJ, XZ, ML, ZW, BL, SY and YY contributed to wastewater sampling point layout and sample collection. XL, JY and SX contributed to epidemiological investigation. YZ, MLC, XS, BP, QC, YQ, SW, MJ, MMC, JT, LW and LH contributed to sample testing and data acquisition. QH, XZ, SF, TZ, HL and CW reviewed and revised the manuscript. All authors approved the final version. All authors had full access to all the data in the study and had final responsibility for the decision to submit for publication.

## Declaration of interests

We declare no competing interests.

## Acknowledgments

This work was supported by the Sanming Project of Medicine in Shenzhen (SZSM201811071) and Shenzhen Key Medical Discipline Construction Fund (SZXK064). We thank the professors of Jianguo Xu, Zijian Feng, Ruifu Yang, Biao Kan, Wenbo Xu, Song Tang, Lan Zhang, and Jianfeng He from Chinese Center for Disease Control and Prevention (JX, BK, WX, ST and LZ), Chinese Preventive Medicine Association (ZF), Academy of Military Medical Sciences (RY) and Guangdong Provincial Center for Disease Control and Prevention (JH) for their professional advising on the establishment of wastewater surveillance system in Shenzhen. We thank Prof. Chao Yang for his guidance on sequencing analysis. We also thank the teams of Shenzhen Hybribio Medical Laboratory and ShenZhen DiAn HuXin Medical Laboratory for their contributions on sample testing.

## Notes

### Competing Interest Statement

The authors have declared no competing interest.

### Funding Statement

This study was funded by the Sanming Project of Medicine in Shenzhen (SZSM201811071) and Shenzhen Key Medical Discipline Construction Fund (SZXK064).

### Author Declarations

Ethics committee of Shenzhen Center for Disease Control and Prevention gave ethical approval for this work

## Reference

1. Crank K, Chen W, Bivins A, Lowry S, Bibby K. Contribution of SARS-CoV-2 RNA shedding routes to RNA loads in wastewater. Sci Total Environ 2022; 806(Pt 2): 150376.

2. Wannigama DL, Amarasiri M, Hurst C, et al. Tracking COVID-19 with wastewater to understand asymptomatic transmission. Int J Infect Dis 2021; 108: 296–9.

3. Lodder W, de Roda Husman AM. SARS-CoV-2 in wastewater: potential health risk, but also data source. Lancet Gastroenterol Hepatol 2020; 5(6): 533–4.

4. Peccia J, Zulli A, Brackney DE, et al. Measurement of SARS-CoV-2 RNA in wastewater tracks community infection dynamics. Nat Biotechnol 2020; 38(10): 1164–7.

5. Wu F, Zhang J, Xiao A, et al. SARS-CoV-2 Titers in Wastewater Are Higher than Expected from Clinically Confirmed Cases. mSystems 2020; 5(4).

6. Medema G, Heijnen L, Elsinga G, Italiaander R, Brouwer A. Presence of SARS-Coronavirus-2 RNA in Sewage and Correlation with Reported COVID-19 Prevalence in the Early Stage of the Epidemic in The Netherlands. Environmental Science & Technology Letters 2020; 7(7): 511–6.

7. Guo M, Tao W, Flavell RA, Zhu S. Potential intestinal infection and faecal-oral transmission of SARS-CoV-2. Nat Rev Gastroenterol Hepatol 2021; 18(4): 269–83.

8. Daughton CG. Wastewater surveillance for population-wide Covid-19: The present and future. Sci Total Environ 2020; 736: 139631.

9. COVIDPoops19 Summary of Global SARS-CoV-2 Wastewater Monitoring Efforts by UC Merced Researchers. 2023. https://ucmerced.maps.arcgis.com/apps/opsdashboard/index.html#/c778145ea5bb4daeb58d31afee 389082 (accessed Feb 28 2023).

10. Bowes DA, Driver EM, Kraberger S, et al. Leveraging an established neighbourhood-level, open access wastewater monitoring network to address public health priorities: a population-based study. Lancet Microbe 2023; 4(1): e29–e37.

11. Monthly statistics for the Environmental Monitoring for Health Protection (EMHP) wastewater programme (England). 2023. https://www.gov.uk/government/collections/monthly-statistics-for-the-environmental-monitoring-for-health-protection-emhp-wastewater-program-england (accessed Feb 28 2023).

12. Jahn K, Dreifuss D, Topolsky I, et al. Early detection and surveillance of SARS-CoV-2 genomic variants in wastewater using COJAC. Nat Microbiol 2022; 7(8): 1151–60.

13. Xu X, Zheng X, Li S, et al. The first case study of wastewater-based epidemiology of COVID-19 in Hong Kong. Sci Total Environ 2021; 790: 148000.

14. Ahmed W, Angel N, Edson J, et al. First confirmed detection of SARS-CoV-2 in untreated wastewater in Australia: A proof of concept for the wastewater surveillance of COVID-19 in the community. Sci Total Environ 2020; 728: 138764.

15. Fu S, He F, Wang R, et al. Development of quantitative wastewater surveillance models facilitated the precise epidemic management of COVID-19. Sci Total Environ 2023; 857(Pt 1): 159357.

16. McMahan CS, Self S, Rennert L, et al. COVID-19 wastewater epidemiology: a model to estimate infected populations. Lancet Planet Health 2021; 5(12): e874–e81.

17. Liu J, Liu M, Liang W. The Dynamic COVID-Zero Strategy in China. China CDC Wkly 2022; 4(4): 74–5.

18. Deng Y, Zheng X, Xu X, et al. Use of Sewage Surveillance for COVID-19: A Large-Scale Evidence-Based Program in Hong Kong. Environ Health Perspect 2022; 130(5): 57008.

19. Zheng X, Wang M, Deng Y, et al. A rapid, high-throughput, and sensitive PEG-precipitation method for SARS-CoV-2 wastewater surveillance. Water Res 2023; 230: 119560.

20. Yang C, Li Y, Jiang M, et al. Outbreak dynamics of foodborne pathogen Vibrio parahaemolyticus over a seventeen year period implies hidden reservoirs. Nat Microbiol 2022; 7(8): 1221–9.

21. Karthikeyan S, Levy JI, De Hoff P, et al. Wastewater sequencing reveals early cryptic SARS-CoV-2 variant transmission. Nature 2022; 609(7925): 101–8.

22. Jassat W, Abdool Karim SS, Ozougwu L, et al. Trends in Cases, Hospitalization and Mortality Related to the Omicron BA.4/BA.5 Sub-Variants in South Africa. Clin Infect Dis 2022.

23. Nyberg T, Ferguson NM, Nash SG, et al. Comparative analysis of the risks of hospitalisation and death associated with SARS-CoV-2 omicron (B.1.1.529) and delta (B.1.617.2) variants in England: a cohort study. Lancet 2022; 399(10332): 1303–12.

24. Zhou JQ, Liu GX, Huang XL, Gan HT. The importance of fecal nucleic acid detection in patients with coronavirus disease (COVID-19): A systematic review and meta-analysis. J Med Virol 2022; 94(6): 2317–30.

25. Gupta S, Parker J, Smits S, Underwood J, Dolwani S. Persistent viral shedding of SARS-CoV-2 in faeces - a rapid review. Colorectal Dis 2020; 22(6): 611–20.

26. Natarajan A, Zlitni S, Brooks EF, et al. Gastrointestinal symptoms and fecal shedding of SARS-CoV-2 RNA suggest prolonged gastrointestinal infection. Med (N Y) 2022; 3(6): 371–87 e9.

27. Zheng S, Fan J, Yu F, et al. Viral load dynamics and disease severity in patients infected with SARS-CoV-2 in Zhejiang province, China, January-March 2020: retrospective cohort study. BMJ 2020; 369: m1443.

28. Wang W, Xu Y, Gao R, et al. Detection of SARS-CoV-2 in Different Types of Clinical Specimens. JAMA 2020; 323(18): 1843–4.

29. Lin W, Xie Z, Li Y, et al. Association between detectable SARS-COV-2 RNA in anal swabs and disease severity in patients with coronavirus disease 2019. J Med Virol 2021; 93(2): 794–802.

30. Parasa S, Desai M, Thoguluva Chandrasekar V, et al. Prevalence of Gastrointestinal Symptoms and Fecal Viral Shedding in Patients With Coronavirus Disease 2019: A Systematic Review and Meta-analysis. JAMA Netw Open 2020; 3(6): e2011335.

31. Wolfel R, Corman VM, Guggemos W, et al. Virological assessment of hospitalized patients with COVID-2019. Nature 2020; 581(7809): 465–9.

32. Han MS, Seong MW, Kim N, et al. Viral RNA Load in Mildly Symptomatic and Asymptomatic Children with COVID-19, Seoul, South Korea. Emerg Infect Dis 2020; 26(10): 2497–9.

33. Lescure FX, Bouadma L, Nguyen D, et al. Clinical and virological data of the first cases of COVID-19 in Europe: a case series. Lancet Infect Dis 2020; 20(6): 697–706.

34. Cao S, Gan Y, Wang C, et al. Post-lockdown SARS-CoV-2 nucleic acid screening in nearly ten million residents of Wuhan, China. Nat Commun 2020; 11(1): 5917.

35. Guangzhou: over 85% of the population infected with SARS-CoV-2. 2023. https://www.cnr.cn/gd/guangdongyaowen/20230118/t20230118_526130010.shtml (accessed Feb 28, 2023).

36. Prasek SM, Pepper IL, Innes GK, et al. Variant-specific SARS-CoV-2 shedding rates in wastewater. Sci Total Environ 2023; 857(Pt 1): 159165.

