## Appendix for "Longitudinal wastewater surveillance addressed public health priorities during the transition from “dynamic COVID-zero” to “opening up” in China: a population-based study"

### Supplementary appendix

#### Contents


| Serial | Details | Page |
|---|---|---|
| Appendix.1 | Fecal sample processing and analysis method details | 1 |
| Appendix.2 | Wastewater sample processing and analysis method details | 1 |
| Appendix.3 | Supplementary Figure 1. Dynamical change of sampling sites during the "dynamic COVID-zero" period. | 2 |
| Appendix.4 | Supplementary Table 1. Location, cover area, population, water flow rate of WWTPs and pump stations after the public health policy was switched to "open-up" in December 2022. | 3 |
| Appendix.5 | Supplementary Figure 2. Daily flow wastewater measurements for WWTPs. | 4 |
| Appendix.6 | Supplementary Figure 3. Faecal viral shedding dynamics of SARS-CoV-2 cases. | 5 |
| Appendix.7 | Supplementary Figure 4. Factors influencing faecal viral shedding of SARS-CoV-2 cases during the acute period | 6 |
| Appendix.8 | Supplementary Figure 5. Procedures after SARS-CoV-2 RNA detected in wastewater during the "dynamic COVID-zero" period. | 7 |
| Appendix.9 | Supplementary Figure 6. Wastewater surveillance played roles in submitting early warming and revealing cryptic community transmission. | 8 |
| Appendix.10 | Supplementary Table 2. Comparison of the number of SARS-CoV-2-infected persons detected by city-wide nucleic acid screening with the predicted number based on viral RNA copy detected in wastewater in a WWTP (WWTP-FT03) of Futian District. | 9 |
| Appendix.11 | Supplementary Figure 7. Sequencing and lineage assignment of SARS-CoV-2 from human oropharyngeal swab samples in Shenzhen, China. | 10 |

**Appendix.1: Fecal sample processing and analysis method details**

Approximately 20 mg faecal samples were collected using sterilized cotton swabs and placed in 3 mL aliquots in SARS-CoV-2 inactivated sampling tubes. After homogeneous mixing, 200 μL of mixed liquor was used to extract nucleic acids using the GeneRotex96 automatic nucleic acid extraction platform (TIANLONG, China). The extracted RNA (50 μL) was stored at -80°C until detection by TaqMan-based RT–qPCR using the novel coronavirus 2019-nCoV nucleic acid detection kit (Hybribio, China) and ABI Prism7500 real-time PCR instrument (Hybribio, China) according to the manufacturer's instructions. A positive faecal sample indicated that at least one target gene (*ORF1ab* or *N*) had a signal under the cut-off cycle threshold (Ct) value of 40. Quantification of positive faecal samples was calculated using the standard curve of *N*-carrying plasmids ranging from $10^1$ to $10^6$ copies $uL^{-1}$; the reaction efficiency was over 90%. Triplicate standard curves were analysed for each new batch of assay reagents and were used to quantify samples as viral genome copies $L^{-1}$. The *N*-carrying plasmid was synthesized by Sangon Biotechnology (Shanghai, China) with an insertion sequence of the *N* gene of SARS-CoV-2 (reference sequence: MN908947.3) cloned into a pUC57 vector, and the resulting plasmid was transformed into competent *E. coli* TOP10 cells.

**Appendix.2: Wastewater sample processing and analysis method details**

Raw wastewater samples were concentrated by a modified "PEG precipitation method".[1] To be specific, approximately 50 mL aliquots of raw wastewater samples were centrifuged at 2,000 × g for 2 minutes at 4°C. Forty milliliter aliquots of supernatant were moved to 50 mL centrifuge tubes with the addition of PEG (10%, w/v) and NaCl (2%, w/v) and shaken vigorously until PEG dissolved completely. The resulting samples were agitated at 180 rpm for 2 hours on an Innova 44 Incubator Shaker Series (New Brunswick, Eppendorf, Hamburg, Germany). Next, they were centrifuged at 4,750 ×g for 30 min on a Sorvall LYNX 4000 Superspeed Centrifuge (Thermo Scientific, Waltham, MA, USA). The supernatant was discarded, and the pellet was used for RNA extraction using automatic nucleic acid extraction platform HBNP-9601A (Hybribio, China) or GENEDIAN EB1000 (DIAN Biotechnology, China). The extracted RNA (50 μL) was stored at -80°C. RT–qPCR detection and quantification were conducted as mentioned above.

**Appendix.3**

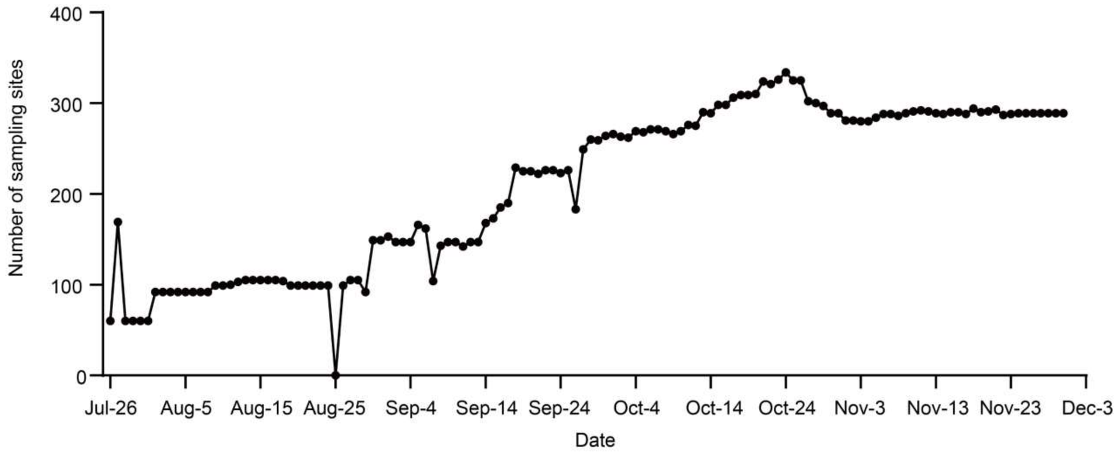


**Supplementary Figure 1.** Dynamical change of sampling sites during the "dynamic COVID-zero" period. From July 26 to October 15, 2022, the number of sampling sites continuously increased from 60 to 298, after which the daily number of sampling sites fluctuated between 280 and 334, with an average of 295 (Figure S1). Classification of sampling sites included residential areas (n=138), urban villages (n=137), port areas (n=15), commercial areas (n=13), farmers' markets (n=7), office buildings (n=2), industrial areas (n=1), nursing homes (n=1), and other types (n=20). The average population covered by each sampling site was approximately 5,200 (50-70,000), and the total population was estimated to be approximately 1·74 million.

**Appendix.4**

**Supplementary Table 1.** Location, cover area, population, water flow rate of WWTPs and pump stations after the public health policy was switched to "open-up" in December 2022.

| **Sampling site** | **Location (longitude and latitude)** | **Cover area (square km)** | **Population** | **Sewage flow ($m^3$/d)** |
|---|---|---|---|---|
| WWTP-FT01 | 114°1′13″E, 22°31′40″N | 21 | 320,000 | 299,000 |
| WWTP-FT02 | 114°0′58″E, 22°31′42″N | 44 | 850,000 | 70,000 |
| WWTP-FT03 | 114°5′47″E, 22°32′8″N | 13 | 450,000 | 97,000 |
| PS-FT01 | 114°4′51″E, 22°31′46″N | 0·75 | 80,000 | 24,000 |
| PS-FT02 | 114°4′35″E, 22°32′28″N | 0·.87 | 12,000 | 6,000 |
| PS-FT03 | 114°3′53″E, 22°31′29″N | 0·36 | 52,000 | 16,000 |
| PS-FT04 | 114°0′52″E, 22°33′37″N | 0·21 | 10,000 | 5,000 |
| PS-FT05 | 114°3′1″E, 22°31′29″N | 0·12 | 26,000 | 5,000 |
| PS-FT06 | 114°3′11″E, 22°30′7″N | 2·3 | 70,000 | 15,000 |
| WWTP-NS01 | 113°53′38″E, 22°27′53″N | 12·5 | 180,000 | 44,000 |
| WWTP-NS02 | 113°53′46″E, 22°30′54″N | 103 | 2,300,000 | 557,000 |
| WWTP-NS03 | 113°57′20″E, 22°35′25″N | 12·8 | 65,000 | 29,000 |
| PS-NS01 | 113°55′51″E,22°31′4″N | 3·865 | 37,000 | 30,000 |
| PS-NS02 | 113°54′49″E,22°31′7″N | 52·68 | 505,000 | 200,000 |
| PS-NS03 | 113°54′32″E,22°31′31″N | 59·66 | 572,000 | 300,000 |

Abbreviation: WWTP, wastewater treatment plant; PS, pump station; FT, Futian District; NS, Nanshan District.

**Appendix.5**

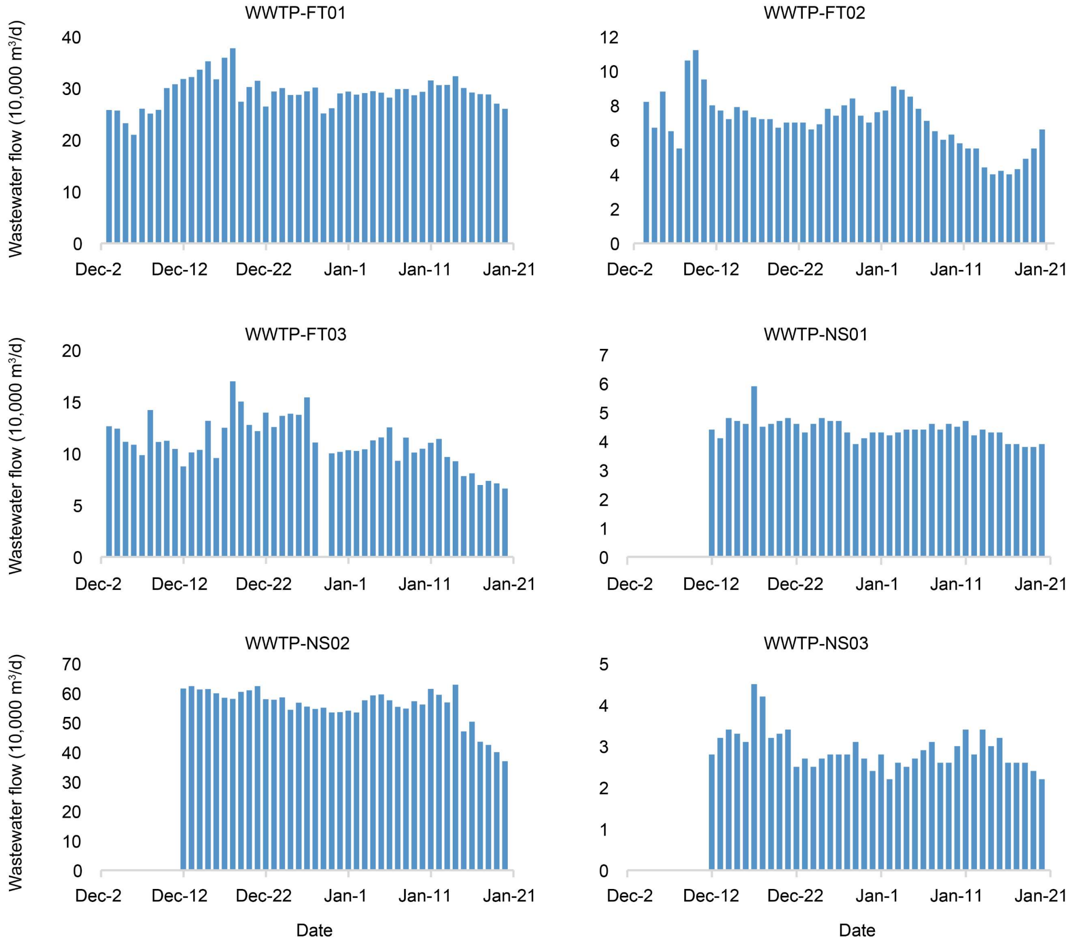


**Supplementary Figure 2.** Daily flow wastewater measurements for WWTPs. Data are only shown for days when samples were collected. WWTP-FT01, WWTP-FT02, and WWTP-FT03 were located in Futian District, and WWTP-NS01, WWTP-NS02, and WWTP-NS03 were located in Nanshan District.

**Appendix.6**

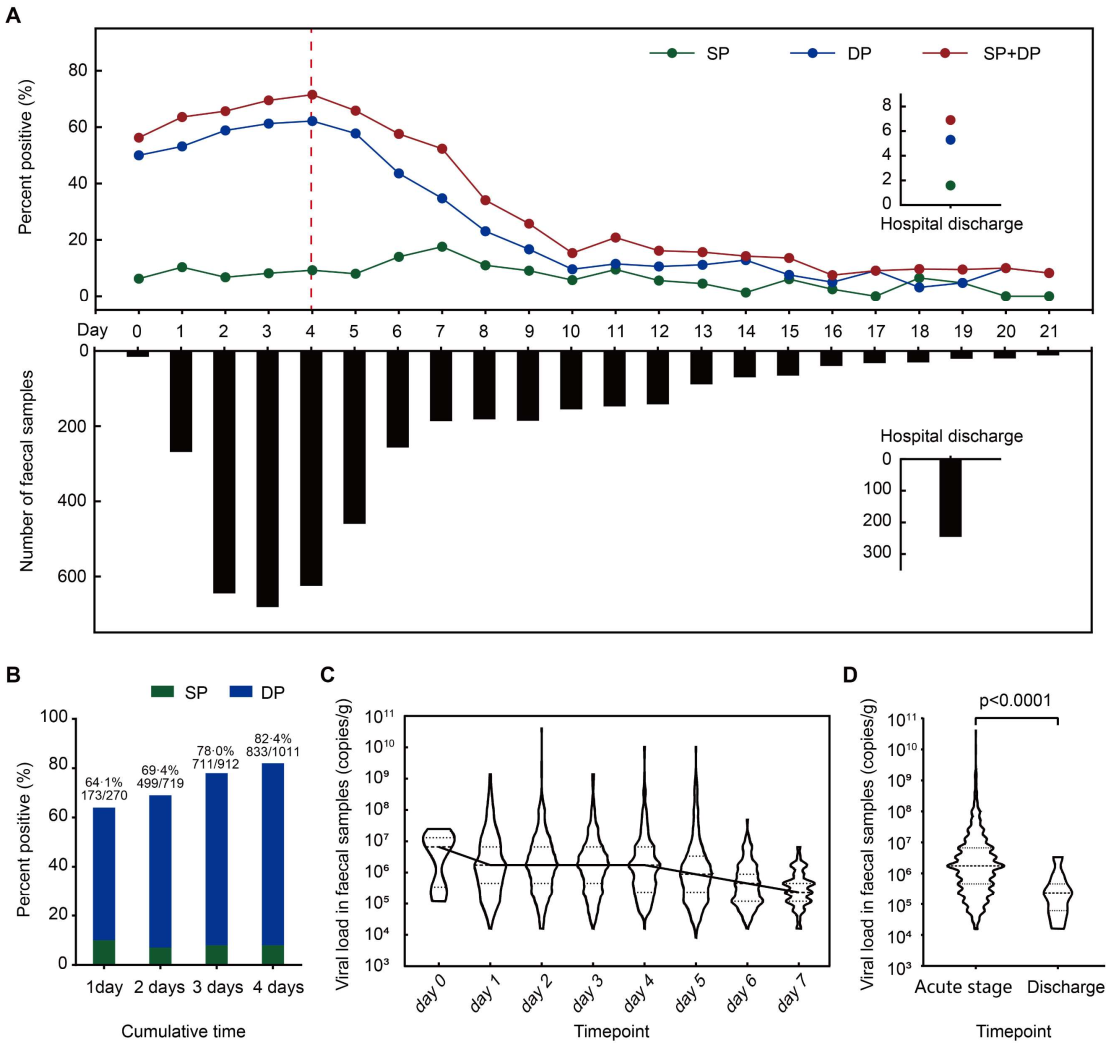


**Supplementary Figure 3.** Faecal viral shedding dynamics of SARS-CoV-2 cases. **(A)** Daily faecal viral shedding proportion of participants. Time was treated since the day when participants tested positive for SARS-CoV-2 RNA after swabbing their throat as a continuous variable. SP, single positive; DP, double positive; **(B)** Pooled faecal viral shedding proportion of participants; **(C)** Faecal viral load dynamics of participants with faecal shedding; **(D)** Faecal viral load comparison between participants during the acute stage and those discharged from the hospital.

**Appendix.7**

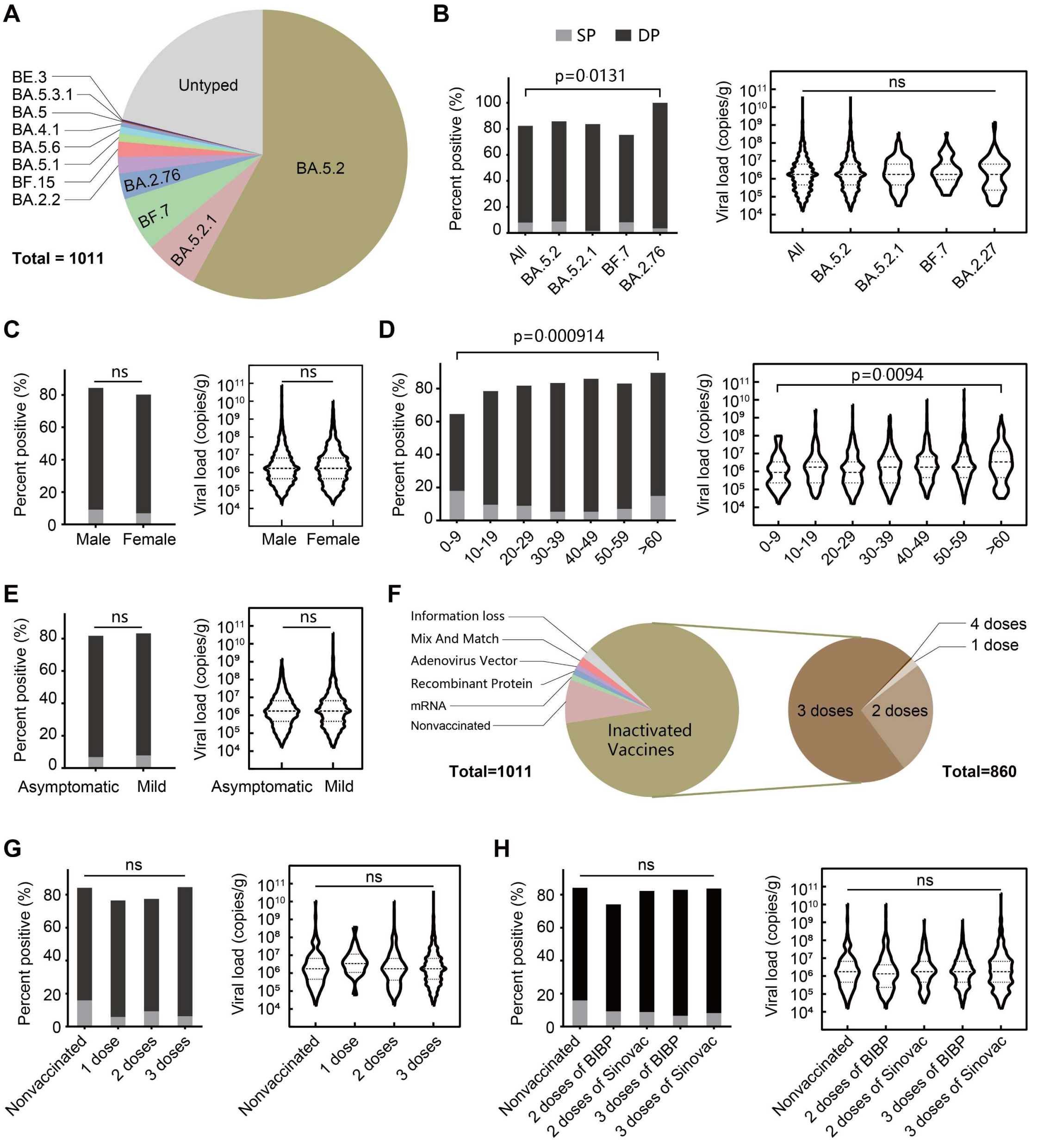


**Supplementary Figure 4.** Factors influencing faecal viral shedding of SARS-CoV-2 cases during the acute period. **(A)** Proportion of viral subvariants of participants during the acute period. The proportion and viral load of participants with faecal shedding were compared between **(B)** Subvariants of Omicron; **(C)** Males and females; **(D)** Different ages; **(E)** Mild cases and asymptomatic carriers. **(F)** Proportion of vaccinations of participants during the acute period. The proportion and viral load of participants with faecal shedding were compared between **(G)** Doses of inactivated vaccine; **(H)** Inactivated vaccine manufacturer BIBP (Beijing Institute of Biological Products Co., Ltd) and Sinovac (Sinovac Life Sciences Co., Ltd). ns, no significant difference (p>0·05).

#### Appendix.8

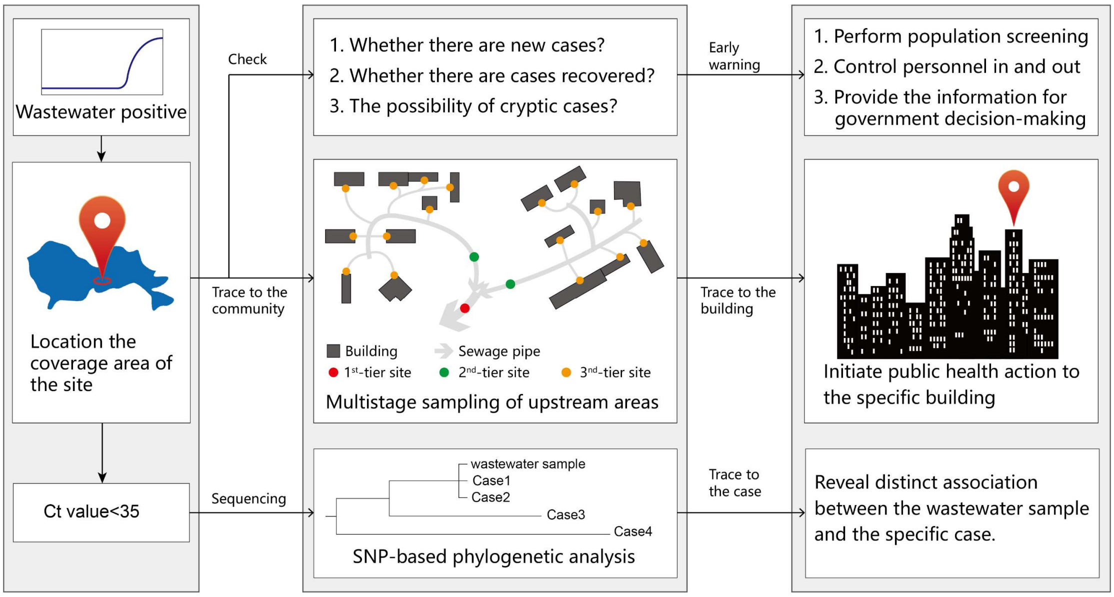


**Supplementary Figure 5.** Procedures after SARS-CoV-2 RNA detected in wastewater during the "dynamic COVID-zero" period.

**Appendix.9**

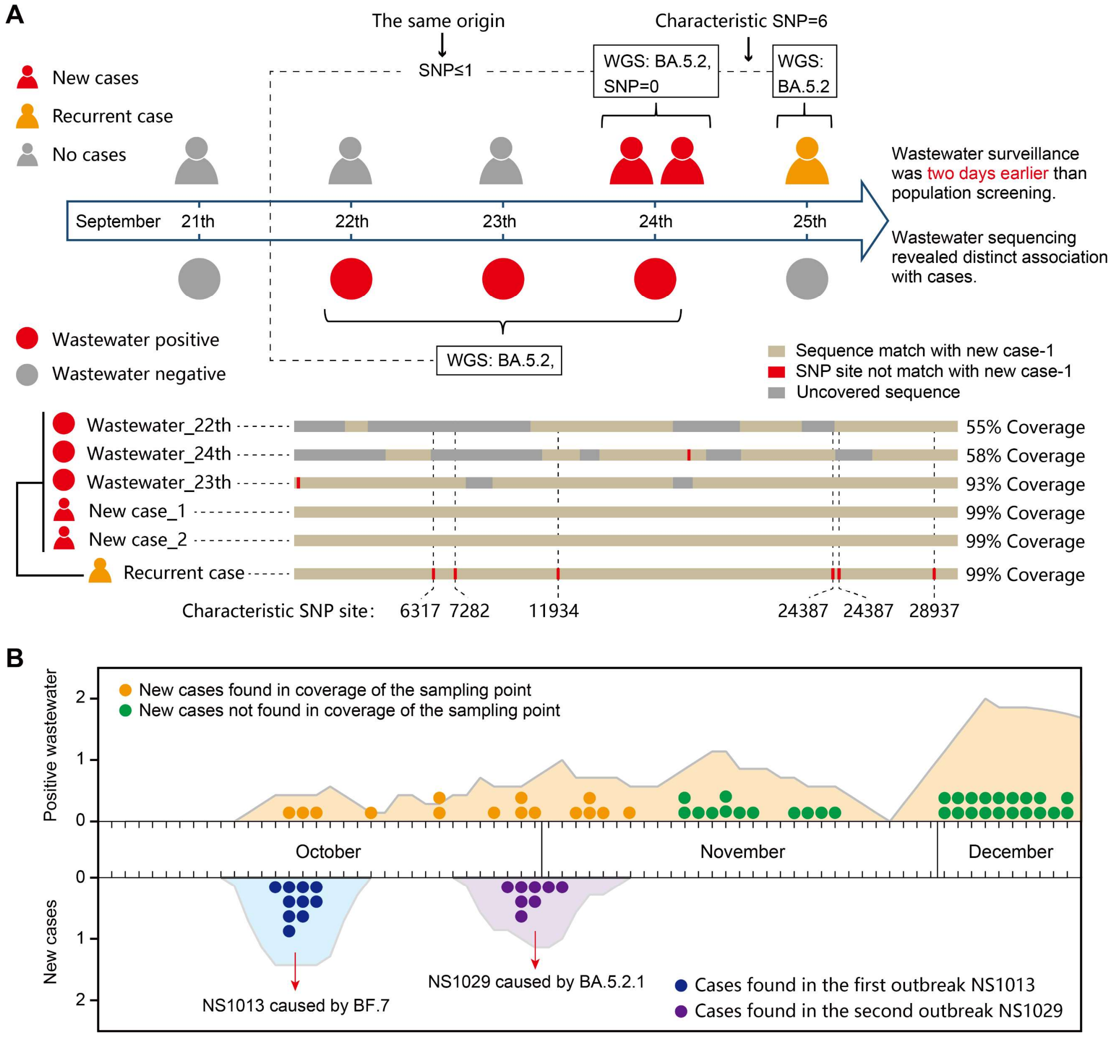


**Supplementary Figure 6.** Wastewater surveillance played roles in submitting early warming and revealing cryptic community transmission. **(A)** Case analysis in Badeng community, wastewater surveillance provided early warming two days earlier than daily city-wide SARS-CoV-2 nucleic acid screening and found that virus RNA in wastewater was derived from two cases identified in September 24 but not from the recurrent case identified in September 25, 2022. **(B)** Case analysis in Nanshan community, after two outbreaks caused by Omicron subvariant BF.7 and BA.5.2.1, respectively, wastewater signal was detected continuously in November, 2022 but no associated cases were identified, suggesting cryptic transmissions in Nanshan community.

**Appendix.10**

**Supplementary Table 2.** Comparison of the number of SARS-CoV-2-infected persons detected by city-wide nucleic acid screening with the predicted number based on viral RNA copy detected in wastewater in a WWTP (WWTP-FT03) of Futian District.

| **Sampling time** | **Infection cases detected by city-wide nucleic acid screening** | **Predicted infection cases median (95% CI)** |
|---|---|---|
| 3-Dec-2022 | 2 | 9·3 (2·33-18·28) |
| 5-Dec-2022 | 22 | 20·11 (7·4-89·6) |
| 6-Dec-2022 | 15 | 16·2 (3·1-37·2) |
| 7-Dec-2022 | 26 | 50·2 (11·5-116·1) |

**Appendix.11**

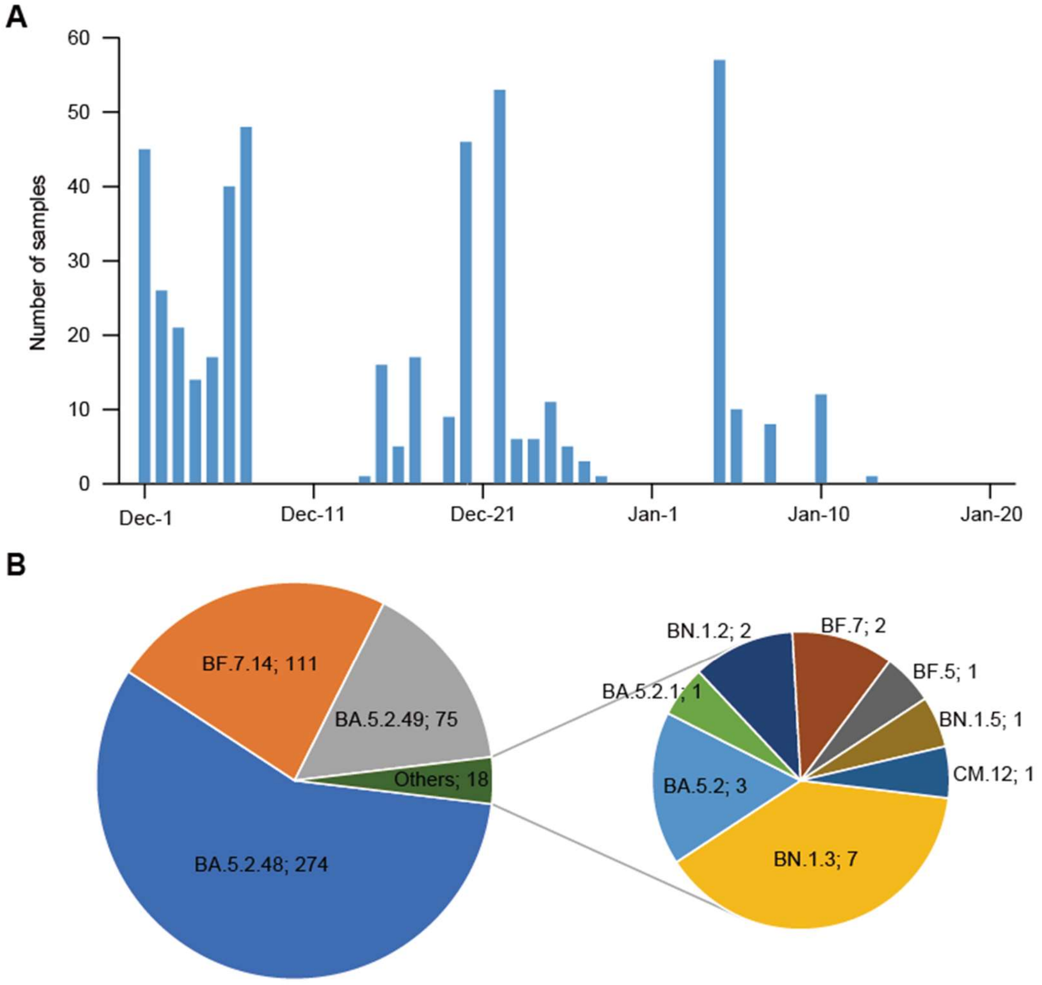


**Supplementary Figure 7.** Sequencing and lineage assignment of SARS-CoV-2 from human oropharyngeal swab samples in Shenzhen, China. **(A)** The number of samples collected during the study period. **(B)** The lineages of SARS-CoV-2 of human oropharyngeal swab samples (n=478).

#### Reference

1. Zheng X, Wang M, Deng Y, et al. A rapid, high-throughput, and sensitive PEG-precipitation method for SARS-CoV-2 wastewater surveillance. *Water Res* 2023; **230**: 119560.
